# Identification of novel type 1 and type 2 diabetes genes by co-localization of human islet eQTL and GWAS variants with *colocRedRibbon*

**DOI:** 10.1101/2024.10.19.24315808

**Authors:** Anthony Piron, Florian Szymczak, Lise Folon, Daniel J. M. Crouch, Theodora Papadopoulou, Maria Inês Alvelos, Maikel L. Colli, Xiaoyan Yi, Marcin Pekalski, Type 2 Diabetes Global Genomics Initiative, Matthieu Defrance, John A. Todd, Décio L. Eizirik, Josep M. Mercader, Miriam Cnop

**Author notes:** https://diagram-consortium.org/T2DGGI.html (Type 2 Diabetes Global Genomics Initiative).

## Abstract

Over 1,000 distinct genetic variants have been associated with diabetes risk by genome-wide association studies (GWAS) but for most their functional impact is unknown and less than 15% of the diabetes GWAS variants have been shown in expression quantitative trait locus (eQTL) studies to alter gene expression in pancreatic islets. To fill this gap, we developed a new co-localization pipeline, called *colocRedRibbon*, that prefilters eQTL variants by direction of effect on gene expression, shortlists overlapping eQTL and GWAS variants and then runs the co-localization. Applying *colocRedRibbon* to diabetes and glycemic trait GWAS, we identified 292 co-localizing gene regions - 236 of which are new - including 24 co-localizations for type 1 diabetes and 268 for type 2 diabetes and glycemic traits. We achieved a four-fold increase in co-localizations, with the novel pipeline and updated GWAS each contributing two-fold. Among the co-localizations are a low frequency variant increasing *MYO5C* expression that reduces type 2 diabetes risk and a type 1 diabetes protective variant that increases *FUT2* and decreases *RASIP1* expression. These novel co-localizations represent a significant step forward to understand polygenic diabetes genetics and its impact on human islet gene expression.

## INTRODUCTION

Diabetes is a complex multi-factorial disease characterized by elevated blood glucose levels. More than 537 million people are affected worldwide, and the prevalence of diabetes is forecasted to reach 1.31 billion by 2050 (1,2). The great majority of patients have type 1 (10-15%) or type 2 diabetes (>85%); secondary or monogenic diabetes is uncommon to rare. Environmental factors such as sedentary lifestyle, obesity, ageing, viral infections, and gut microbiome play an important role in the development of diabetes and underlie the increasing prevalence rates (3–5). Familial clustering and heritability studies have long provided evidence for the role of genetic factors and type 1 and type 2 diabetes are now established as polygenic. Progressive pancreatic islet dysfunction, caused by a combination of environmental and genetic factors, is central in most forms of diabetes (6–8). Therefore, the study of human islet pathophysiology is essential to better understand the disease.

Large genome-wide association studies (GWAS) and fine-mapping analyses have associated more than 700 genetic loci to risk for type 1 and type 2 diabetes and glycemic traits (9–15), but the mechanism of action for most remains poorly understood. 89% of the fine-mapped type 2 diabetes GWAS variants lie in non-coding regions in the genome (9); these are typically named after the closest gene. Cis-expression quantitative trait locus (cis-eQTL) analyses link variants with gene expression. Such approaches have identified gene expression changes associated with variants lying farther away or in intronic regions of other genes in many tissues or cells (16) and in human islets (17,18), showing that the closest gene is not necessarily the effector gene. eQTLs further inform about direction of effect, i.e. whether a variant is associated with lower or higher gene expression. Linking diabetes-associated variants to gene expression is therefore an essential step to understand diabetes genetics and pathophysiology.

One of the methods to link disease variants with gene expression is to co-localize GWAS and cis-eQTL variants. The Genotype-Tissue Expression (GTEx) project generated eQTL analyses for 54 human tissues (19), among which pancreas. Human islets - a key tissue in diabetes pathogenesis - are not part of the GTEx tissue collection. Separate initiatives have been developed to identify human islet eQTLs and co-localize them with diabetes GWAS hits, making progress in the domain, but falling short of expectations: only around 50 are associated with specific gene expression variation (17,18). We hypothesized that this missing link is, at least in part, caused by methodological limitations of the available co-localization approaches. Indeed, we observed that existing co-localization tools miss matching GWAS and eQTL signals when analyzing chromosomal regions that contain strong multiple distinct GWAS signals. These tools also do not consider the direction of effect of variants in a region tested for colocalization.

To test this hypothesis and fill the co-localization gap, we developed a new co-localization pipeline, *colocRedRibbon*. The method uses novel pre-filtering steps, shortlisting variants before running an existing co-localization tool. We use *colocRedRibbon* to co-localize human islet cis-eQTLs (17) and GWAS for type 1 (20), type 2 diabetes (15) and glycemic traits (11). We significantly expand the number of co-localizations for type 2 diabetes with 268 co-localizing gene regions, and detect 24 type 1 diabetes colocalizations, making this study the largest to date and improving knowledge about genetic regulation in human islets.

## MATERIALS AND METHODS

The novel *colocRedRibbon* R package integrates the *coloc* co-localization package (21) with a variant shortlisting method. The shortlisting method is based on *RedRibbon* rank-rank hypergeometric overlap (22) and uses the direction of effect of *GWAS* risk alleles in the eQTL analysis and is described in more detail below.

### colocRedRibbon workflow

*colocRedRibbon* is a tool designed to identify common causal candidates by colocalizing GWAS and eQTL (Figure 1A). This approach aims to pinpoint variants associated both with diabetes risk and gene expression. The method employs a two-step approach for shortlisting variants: a risk allele effect step and a *RedRibbon* overlap step. The risk allele effect step categorizes variants into two disjoint sets based on the direction of effect on gene expression of the disease-risk increasing allele, i.e., down- or upregulating. The upregulating variant set comprises the variants whose risk alleles increase gene expression, and the downregulating set risk alleles that decrease gene expression. The subsequent steps are applied independently on both variant sets.

**Figure 1.**
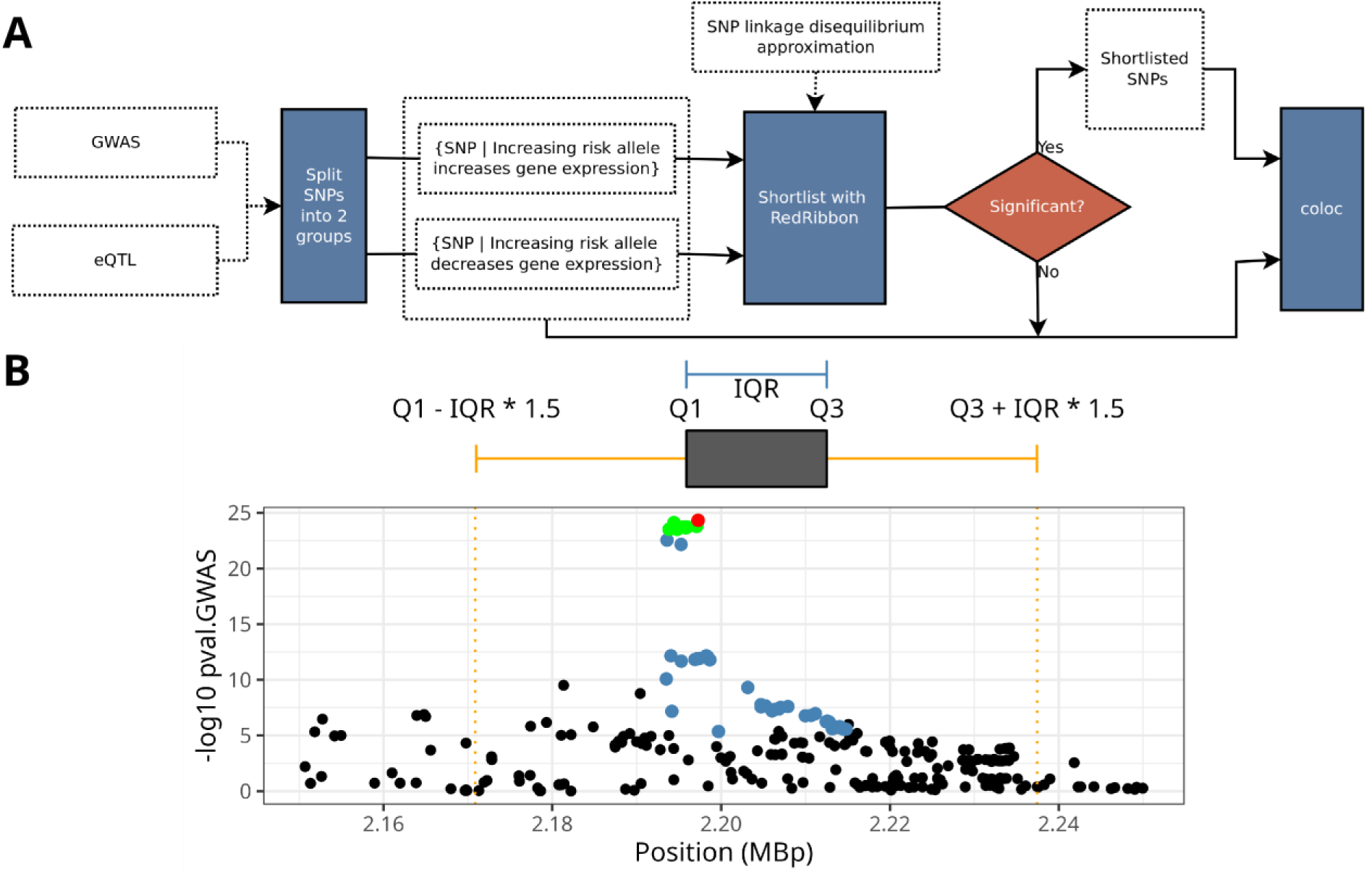
The *colocRedRibbon* pipeline. (A) The workflow of *colocRedRibbon*. GWAS and eQTL SNPs are split into two effect sets depending on the direction of effect on gene expression for the increasing risk allele. *RedRibbon* rank-rank hypergeometric overlap method is applied on both SNP sets ranked by the P-values. If a significant overlap is detected by *RedRibbon*, these shortlisted SNPs are analyzed by *coloc*. Otherwise, *coloc* is applied to the two effect sets without overlap shortlisting. (B) The interquartile range mode of *colocRedRibbon* adds a background of variants between the dotted orange lines where *Q*_1_is the 25^th^ percentile (first quartile) of the chromosomal positions of the core set, *Q*_2_the 75^th^ percentile (third quartile), and *IQR* is the interquartile range.

In the shortlisting step, the *RedRibbon* rank-rank hypergeometric overlap method (22) is applied on GWAS and eQTL variants. This step involves the ranking of variants by *P*-values for both GWAS or eQTLs and statistically examining potential overlaps between the ranked lists. If significant overlap is detected, the shortlisted SNPs are further analyzed by the *coloc* package. In cases where *RedRibbon* overlap is not significant, *coloc* analysis is performed without prior shortlisting.

### colocRedRibbon shortlisting

The shortlisting step leverages a known biological principle to categorize variants into two disjoint subsets. This principle is based on the expectation that risk alleles of multiple variants within a gene region that are in linkage disequilibrium should produce concordant expression effects, either upregulating or downregulating gene expression. Risk alleles of variants are hence split into two sets, one upregulating and the other downregulating a gene. Only one of these two sets is likely to yield meaningful co-localization results, as relevant associations are expected to have consistent directional effects. Compared to analyses that do not include this step, this division typically halves the number of candidate loci, streamlining subsequent analyses.

#### RedRibbon overlap shortlisting

The *RedRibbon* method (22) ranks GWAS and eQTL variant lists by increasing *P*-value, prioritizing variants with the strongest statical significance. *RedRibbon* aims to uncover the best overlap among variants with the lowest *P*-values in both lists. For each coordinate (*i*, *j*) in the two lists, where *i* represents the GWAS rank and *j* the eQTL rank, an overlap *P*-value is computed using a one-sided hypergeometric statistic. Let *c* be the number of variants common to both lists at the coordinate (*i*, *j*) in a *RedRibbon* overlap map of size *n* × *n*, where *n* is the number of elements in each list, the overlap *P-value* is calculated as:

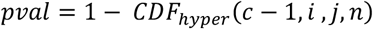

where *CDF*_*hyper*_ represents the hypergeometric cumulative distribution function. This *P-value* quantifies the probability of observing *c* or more overlapping variants by chance, given the positions in the ranked lists. *RedRibbon* overlap results in a list of variants corresponding to the minimal *P*-value overlap. This approach allows for a statistical assessment of the overlap between GWAS and eQTL SNPs, facilitating the identification of potential causal variants that influence both disease risk and gene expression.

#### RedRibbon overlaps shortlisting in interquartile range mode

The list of shortlisted variants can be directly fed into the *coloc* tool. Or, alternatively, the interquartile range (IQR) method can be employed to add background, providing a more comprehensive analysis (Figure 1B). The RedRibbon-shortlisted variants serve as the core set. The core set is expanded with additional variants based on their chromosomal positions, using as lower bound, *Q*_1_ − 1.5 ∗ *IQR* and upper bound *Q*_2_ + 1.5 ∗ *IQR* where *Q*_1_ is the 25^th^ percentile (first quartile) of the chromosomal positions of the core set, *Q*_2_the 75^th^ percentile (third quartile), and *IQR* is the interquartile range (*Q*3 − *Q*1). This method adds a background of variants while excluding potential outliers, providing a broader genomic context and ensuring a more robust set for *coloc* analysis.

#### Adjusted P-value computation

The *colocRedRibbon* adjusted minimal *P*-value is computed, leveraging the *P*-value adjustment capabilities of the *RedRibbon* package (22). This approach utilizes a hybrid permutation-prediction method that splits the variants into two sets, the permuted and predicted set. The permuted set undergoes, as the name indicates, random permutation. The predicted set is computed from the permuted set values with the following:

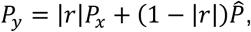

where the *P*-value *P*_*y*_ for variant *y* is predicted from the *P*-value *P*_*x*_ for variant x, the coefficient of correlation *r* representing the linkage disequilibrium, and a random *P*-value ^*P*^^ generated from the distribution of all values in the original list.

The hybrid permutation-prediction of *colocRedRibbon* uses a linkage disequilibrium approximation to compute the adjusted *P*-value. This approximation is given by *r* function, an approximation of the coefficient of correlation between two variants:

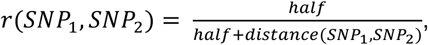

where *distance* is the distance in base pairs between the variants under consideration, and *half* the distance between the variants where *r* equals 0.5.

The rationale behind the approximation is to trade-off some of the adjusted *P*-value accuracy for performance, because using real linkage disequilibrium would be computationally expensive as it needs to be computed for all pairs of variants. Subsequent *coloc* analysis is expected to filter out the remaining false positives.

### colocRedRibbon R package

*colocRedRibbon* is available as a user-friendly R package (see data availability section), designed to streamline the process of colocalization analysis. This package seamlessly combines risk allele effect and *RedRibbon* shortlisting steps with the *coloc* package (21), providing a comprehensive analysis pipeline. It leverages the fast and accurate rank-rank overlap method from *RedRibbon* R package (22). The speed of *RedRibbon* allows for thousands of co-localizations to be run within hours on standard computer hardware.

### Pathway and regulatory elements enrichment

Pathway enrichment was done with *gprofiler2* (v0.1.9) and regulatory element enrichment with the hypergeometric model using Ensembl Regulatory Build GRCh37 (23) and human islet chromatin regions from (24,25). We used *fGSEA* R package (26) to overlap the co-localizing genes with type 2 diabetes differential gene expression from (27).

### Normalization of gene expression

In the gene expression plots, expression was normalized to the geometric mean of stable genes *VAPA*, *ACTB* and *ARF1* (in transcripts per million) (28). For each sample numbered *j*, an adjustment factor *β*_*j*_ is computed with

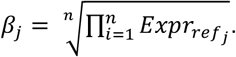

The expression adjustment is computed for each sample *j* and gene *i* as *AdjExpr*_*i*,*j*_ = *β*_*j*_ ∗ *Expr*_*i*_. This normalization was used only for the purpose of visualization of gene expression.

## RESULTS

Previous studies have linked diabetes GWAS signals to eQTL variants in human islets, a highly disease-relevant tissue that hosts the endocrine cells maintaining blood glucose homeostasis. The yield of the 2 largest studies to date (17,18) has been modest at best, identifying co-localization with eQTL variants for only 7% of the current GWAS hits. To investigate the hypothesis that methodological limitations in co-localization approaches contributed to the “co-localization missing link”, we applied the novel *colocRedRibbon* tool to recent large-scale eQTL (17) and GWAS datasets (11,15,20). The eQTL dataset is from the Translational human pancreatic Islet Genotype tissue-Expression Resource (TIGER), which is, with its meta-analysis of eQTLs in 4 cohorts comprising altogether 404 human islet samples, one of the largest eQTL analyses of human islets (17). The European and multi-ancestry type 2 diabetes GWAS datasets were obtained from the Type 2 Diabetes Global Genomic Initiative (T2DGGI) (15). The Wellcome Centre for Human Genetics provided the GWAS for type 1 diabetes (20). The quantitative glycemic traits GWAS originate from MAGIC (11). These GWAS are among the biggest to date, having included >2.5M cases/controls for type 2 diabetes, >170K for type 1 diabetes, and up to) 200K for glycemic traits (Supplementary Table S7).

### Co-localizing variants identifies important diabetes genes

With this newly developed *colocRedRibbon* pipeline, we identified a total of 434 co-localizations between eQTL and GWAS variants. These co-localizations are shown by their chromosomal position in a circle plot, highlighting the regulated genes for GWAS SNPs for type 1 diabetes (n=24), type 2 diabetes (multi-ancestry GWAS n=216; European ancestry n=143) and the glycemic traits fasting glucose (n=22), post-challenge 2-hour glucose (n=4), HbA1c (n=20) and fasting insulin (n=2) (Figure 2, Supplementary Tables S1 and S4).

**Figure 2.**
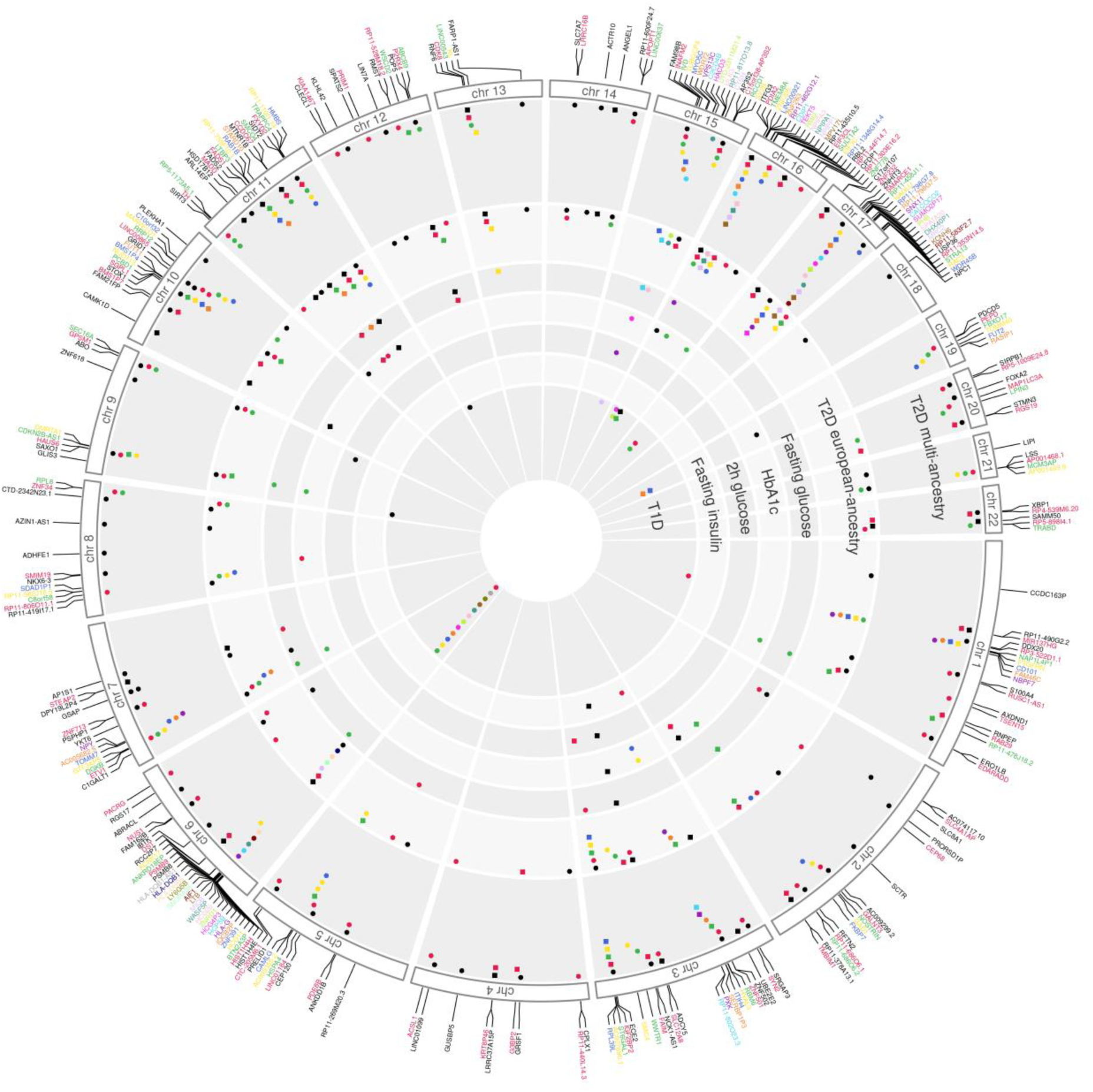
Circle plot of 434 *colocRedRibbon* co-localizations of human islet eQTLs with diabetes and glycemic trait GWAS. eQTL genes are positioned at their starting position on the chromosome. The concentric circles correspond to GWAS. Each dot corresponds to a SNP and the color matches the eQTL gene color. Square dots indicate identical SNPs. (T1D: type 1 diabetes, T2D : type 2 diabetes.)

The co-localization analysis examined a total of 10,835,771 GWAS variants, including non-significant ones, within a 2 million base pair region around the transcription start site of genes exhibiting at least one significantly associated GWAS variant and one eQTL. The distribution of these 10,835,771 variants across different traits revealed that type 2 diabetes accounts for the majority (73%) of these variants, with multi-ancestry GWAS contributing 41% and European ancestry 32%. Type 1 diabetes represented 18% of the variants, and glycemic traits showed lesser degrees of representation: HbA1c (5%), 2-hour glucose (0.5%), fasting glucose (3%), and fasting insulin (0.5%) (see Supplementary Table S6).

Type 2 diabetes demonstrated the highest number of significant co-localizations (Supplementary Tables S1, S4 and S5). The lowest number of co-localizations was found for fasting insulin, which primarily reflects insulin resistance rather than beta cell function. This aligns with the expectation that insulin resistance would primarily manifest in tissues such as skeletal muscle, liver, or adipose tissue rather than in pancreatic islets.

### *The colocRedRibbon* shortlisting steps solve the issue of multiple GWAS signals

Cis-eQTL signals for a gene typically manifest themselves as a single peak of variants in linkage disequilibrium, whereas GWAS signals around the same gene often comprise multiple independent peaks. This multiplicity of signals can confound *coloc*, potentially leaving legitimate co-localizations undetected. A prime example is observed in a 2 million base pair region surrounding *CCDC67* gene (also known as *DEUP1*). Here, *coloc* recognizes a strong type 2 diabetes GWAS signal and identifies the lead variant (red dot, Figure 3A, top panel) in this signal. However, it overlooks an adjacent, highly significant - albeit weaker - signal (Figure 3A, top panel) that holds potential for a legitimate co-localization given the presence of a strong matching eQTL signal at the same chromosomal position (Figure 3A, bottom panel).

**Figure 3.**
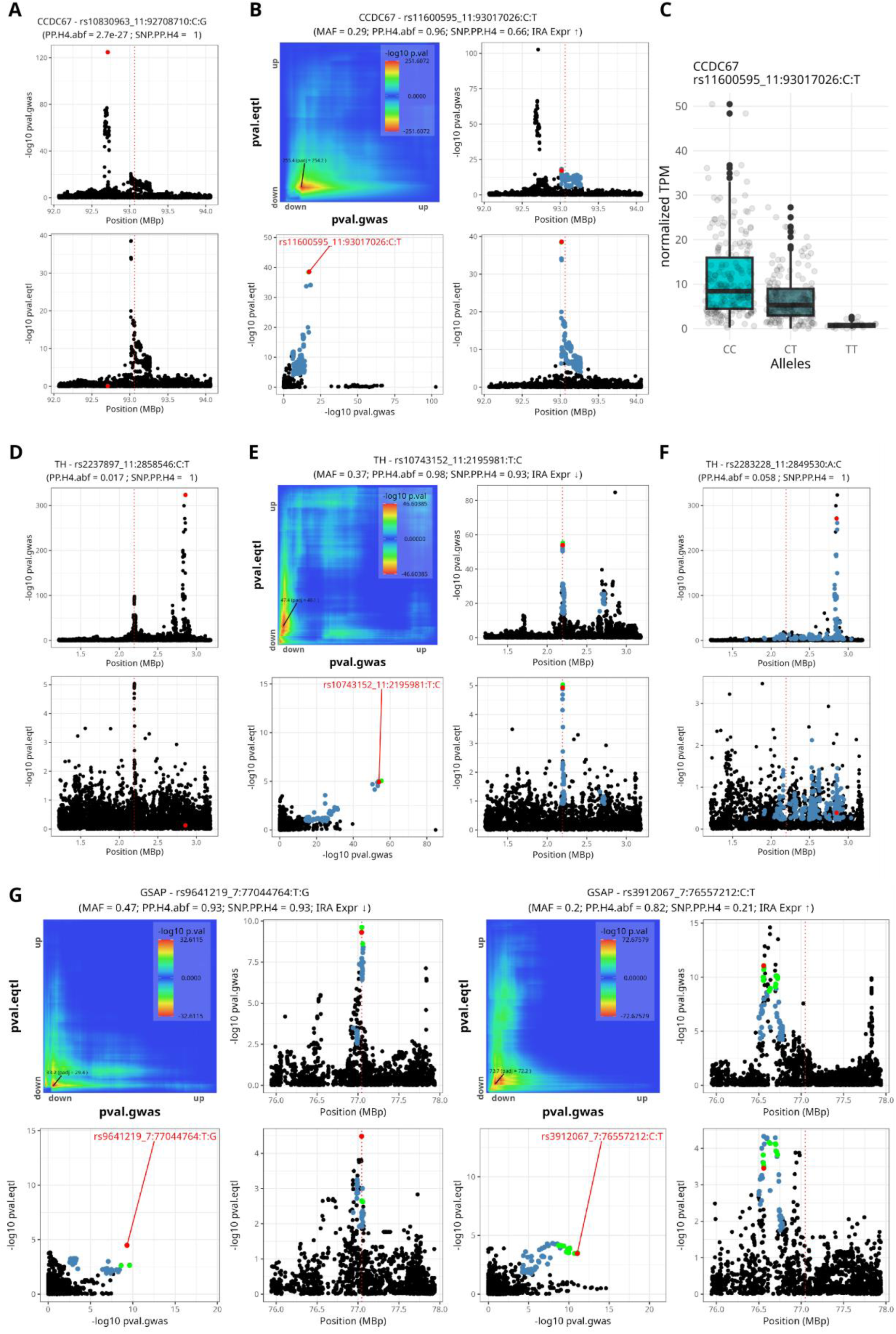
The *colocRedRibbon* shortlisting steps identify genuine co-localizations. (A) GWAS and eQTL Manhattan plots for the *CCDC67* gene region. Two signals are present for GWAS. *coloc* alone does not detect a significant co-localization. The lead SNP (red dot) on the highest GWAS peak does not have an eQTL signal, while the matching eQTL and GWAS peaks are missed by *coloc*. The red dotted line is the gene transcription start site. (B) The *colocRedRibbon* shortlisting method applied to the *CCDC67* gene region detects a colocalization for the lower GWAS signal. The overlapping SNPs — the shortlisted SNPs — are shown in blue and the lead SNP in red in the lower panel. (C) CCDC67 expression according to C or T alleles of the lead SNP identified by *colocRedRibbon*. (D) GWAS and eQTL Manhattan plots for the *TH* gene region. *coloc* does not detect a significant co-localization for the eQTL peak and 6 GWAS peaks. (E) *colocRedRibbon* shortlisting of SNPs that increase diabetes risk (Increasing Risk Allele, IRA) and lower *TH* expression unmasks a co-localization. The most significant GWAS peak (see D) is dismantled by shortlisting of direction of effect of gene expression. The overlap map in the top left panel shows a significant overlap between GWAS and eQTL SNPs. These shortlisted SNPs are shown in color (bottom left): the overlapping SNPs are shown in blue, the 99% credible set in green and the lead SNP in red. (F) There is no colocalization for risk alleles that decrease *TH* gene expression. (G) Co-localization plots for *GSAP* eQTL and type 2 diabetes GWAS. The lead SNP is shown in red, the 99% credible set in green and *RedRibbon* overlap in blue. *GSAP* presents two distinct co-localizations in the same region with opposite risk allele expression regulation. SNPs are referenced by rsids, PP.H4 abf and SNP PP.H4 are the posterior probability for *coloc* tool hypothesis 4, MAF is the minor allele frequency. Arrows pointing up or down indicate increased or reduced gene expression, respectively.

To address this issue of multiple GWAS signals, we shortlisted SNPs using *RedRibbon* overlap. This approach involves ranking GWAS and eQTL variant lists from lowest to highest P-value and searching for overlapping SNPs using a rank-rank hypergeometric overlap approach. The resulting overlap map reveals a strong and highly significant overlap among low *P*-values (Figure 3B, top left panel). By selecting the variants that constitute the most significant overlap (Figure 3B, blue dots in bottom left panel), we successfully identified matching GWAS and eQTL peaks and the lead SNP (Figure 3B, right panels, red dot). This shortlisting step eliminates spurious GWAS signals unrelated to eQTL peaks, enabling *coloc* to detect co-localization using overlapping variants. The identified lead SNP impacts human islet *CCDC67* expression: the *T* effector allele that is associated with reduced type 2 diabetes risk downregulates *CCDC67* transcript expression in human islets (Figure 3C).

The *colocRedRibbon* method further refines the analysis through a direction of effect step, which splits variants into two disjoint subsets based on whether their risk alleles up- or downregulate a gene. This premise reduces the number of variants considered by the *coloc* tool. For instance, in a region around the gene *TH* with multiple type 2 diabetes GWAS signals (Figure 3D, top panel), *coloc* alone fails to detect a co-localization due to the confounding effect of the strongest GWAS signal. By only retaining the variants for which the risk allele downregulates *TH* expression and excluding many highly significant GWAS variants, *coloc* detects another weaker signal (Figure 3E). Conversely, variants for which the risk allele upregulates *TH* expression do not exhibit co-localization (Figure 3F), aligning with the biologically plausible assumption that risk alleles only associate with one direction of effect.

Unexpectedly, five genes colocalize for risk allele expression in both down-regulation and up-regulation. Among these genes is Gamma-Secretase Activating Protein *(GSAP*). *colocRedRibbon* detects two distinct signals going in opposite gene expression direction, while both increase diabetes risk (Figure 3G and Supplementary Table S3). This gamma-secretase catalyzes the intermembrane cleavage of membrane proteins. GSAP plays a role in lipid homeostasis and mitochondrial function (29).

This refined approach demonstrates the power of *colocRedRibbon* in uncovering previously undetected co-localizations and providing more nuanced insights into the relationship between genetic variants, gene expression, and disease risk.

### *colocRedRibbon* doubles the number of co-localizations

The 434 co-localizations between human islet cis-eQTLs and GWAS for type 1 and type 2 diabetes and glycemic traits encompass 344 distinct lead variants, mapping to 289 unique genes (Figure 4, Supplementary Table S4). Only 214 were detected by running *coloc* without the shortlisting steps (Figure 4A); in other words, *colocRedRibbon* doubled detection. Improvements were observed for most GWAS, and the largest was for type 1 diabetes. A total of 307 colocalizations were identified across 236 distinct gene regions for which no co-localization was previously reported.

**Figure 4.**
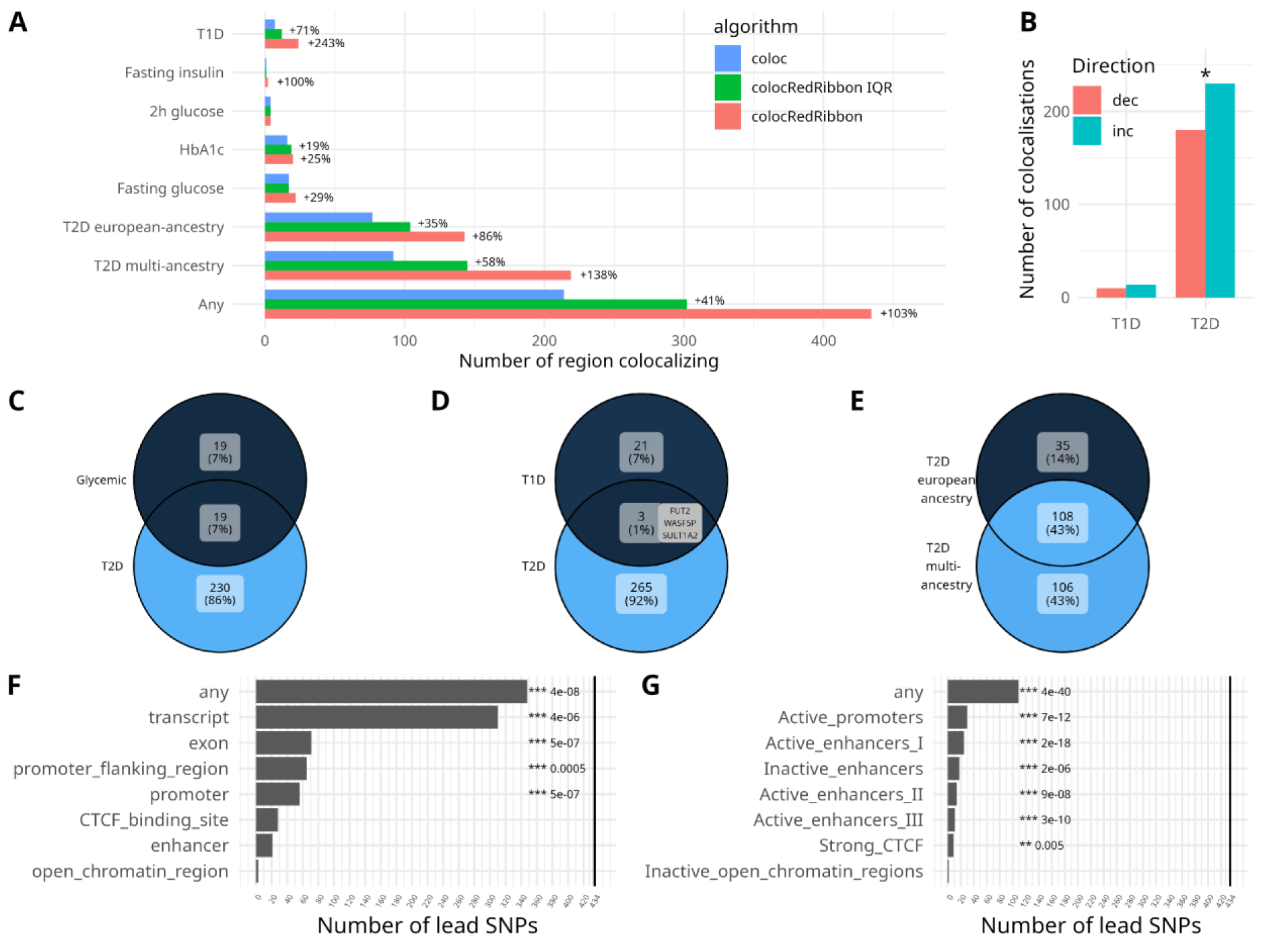
*colocRedRibbon* doubles the number of co-localizations and detected variants are located in islet specific regulatory regions. (A) Number of co-localizing regions. *colocRedRibbon* (red) significantly improves detection of co-localizations compared with *coloc* (blue). The more conservative *colocRedRibbon* interquartile range method (IQR, green) is also better than *coloc*. The percentage increase is shown for GWAS hits for European and multi-ancestry type 2 diabetes (T2D), type 1 diabetes (T1D) and glycemic traits HbA1c, fasting glucose, 2-hour glucose post-challenge, and fasting insulin. “Any” includes all GWAS SNPs combined. The overall detected co-localizations increased by 103%. (B) The direction of effect of the risk allele on gene expression. Inc denotes increased, dec decreased gene expression. (C-E) The number of regulated genes in common for glycemic traits and diabetes GWAS. (F) The enrichment in regulatory regions for lead SNPs. “Any” means any of the regions. (G) Human islet specific regulome enrichment (24,25). *** *P*-Value ≤ 0.001, ** *P*-Value ≤ 0.01, * *P*-Value ≤ 0.05

Out of the 434 identified co-localizations, 48 had a unique lead variant in the 99% credible set (SNP.PP.H4 ≥0.99), and 37% had ≤5 candidate variants in the 99% credible set. This restricted number of credible sets narrows to potential candidates for experimental validation. For instance, in this study, a unique co-localizing lead variant, rs10830963, was detected in *MTNR1B* (Supplementary Table S1). This SNP has been extensively studied and is associated with increased type 2 diabetes risk in some populations (30,31).

For type 2 diabetes co-localizations, risk alleles are significantly more associated with increased gene expression (Figure 4B). Half of the glycemic trait genes are also associated with type 2 diabetes (Figure 4C). Type 1 and type 2 diabetes share only 3 genes (FUT2, SULT1A2, and WASF5P, Figure 4D) with differing alleles and effects on gene expression (See Supplementary Table S1). For type 2 diabetes, 35 genes are specific to European ancestry (Figure 4E and Supplementary Table S2) and 106 are specific to multi-ancestry GWAS. This is not unexpected, as the multi-ancestry GWAS encompasses a broader range of samples, including those of European ancestry.

### Co-localizing variants largely lie in regulatory regions

Co-localizing lead variants showed a notable over-representation in regulatory regions of the genome (Figure 4F, Supplementary Table S8), with 383 lead variants situated in such areas. Significant enrichment was observed in exonic (n=71), promoter (n=56), promoter-flanking (n=65) and transcript regions (n=310). In this tissue-agnostic analysis, no enrichment was detected in open chromatin, enhancer or CTCF-binding sites from ensembl GRCh37 regulatory features (23).

Given the tissue- or cell type-specific nature of many regulatory regions, we further investigated the distribution of co-localizing lead variants within regions known to modulate human islet chromatin structure (25). This analysis revealed a significant enrichment in the human islet regulome (Figure 4G, Supplementary Table S10), particularly in active promoters (n=30), active enhancers (n=50), inactive enhancers (n=18) and strong CTCF-binding regions (n=29). No enrichment was observed in open inactive chromatin regions. When applying the enhancer subclassification by Miguel-Escalada et al., the most significant enrichment of co-localizing lead variants was detected in active class I enhancers (P-value = 2.16 × 10^-18^; n=25), which are characterized by higher H3K27ac marks and stronger Mediator protein occupancy, both key factors in gene expression regulation (25).

In summary, 361 co-localizing lead variants were found to reside in exons, transcripts, regulatory and islet-specific chromatin interaction regions that consist of active promoters and enhancers (23,25). These findings underscore the potential functional importance of these variants in gene regulation and their relevance to disease susceptibility.

### Co-localizing variants identify important pathways

*Gprofiler2* pathway enrichment of pooled co-localizing genes for type 2 diabetes and glycemic traits revealed pathways related to cellular localization, vesicle dynamics, mitochondrial function, fatty acid metabolism and synthesis (Figure 5), suggesting a role for these islet cell processes in the pathogenesis of type 2 diabetes. Notably, genes down-regulated by lead variants were mainly involved in pathways related to cellular localization and trafficking whereas up-regulated genes were primarily associated with cytoplasmic function (Figure 5B-C). Additionally, the co-localizing gene set exhibited significant (*P*-value = 0.00783) overlap with genes differentially expressed in islets from type 2 diabetic vs non-diabetic donors, reported by Marselli et al. (27) (Supplementary Table S1; column fGSEA.t2d).

**Figure 5.**
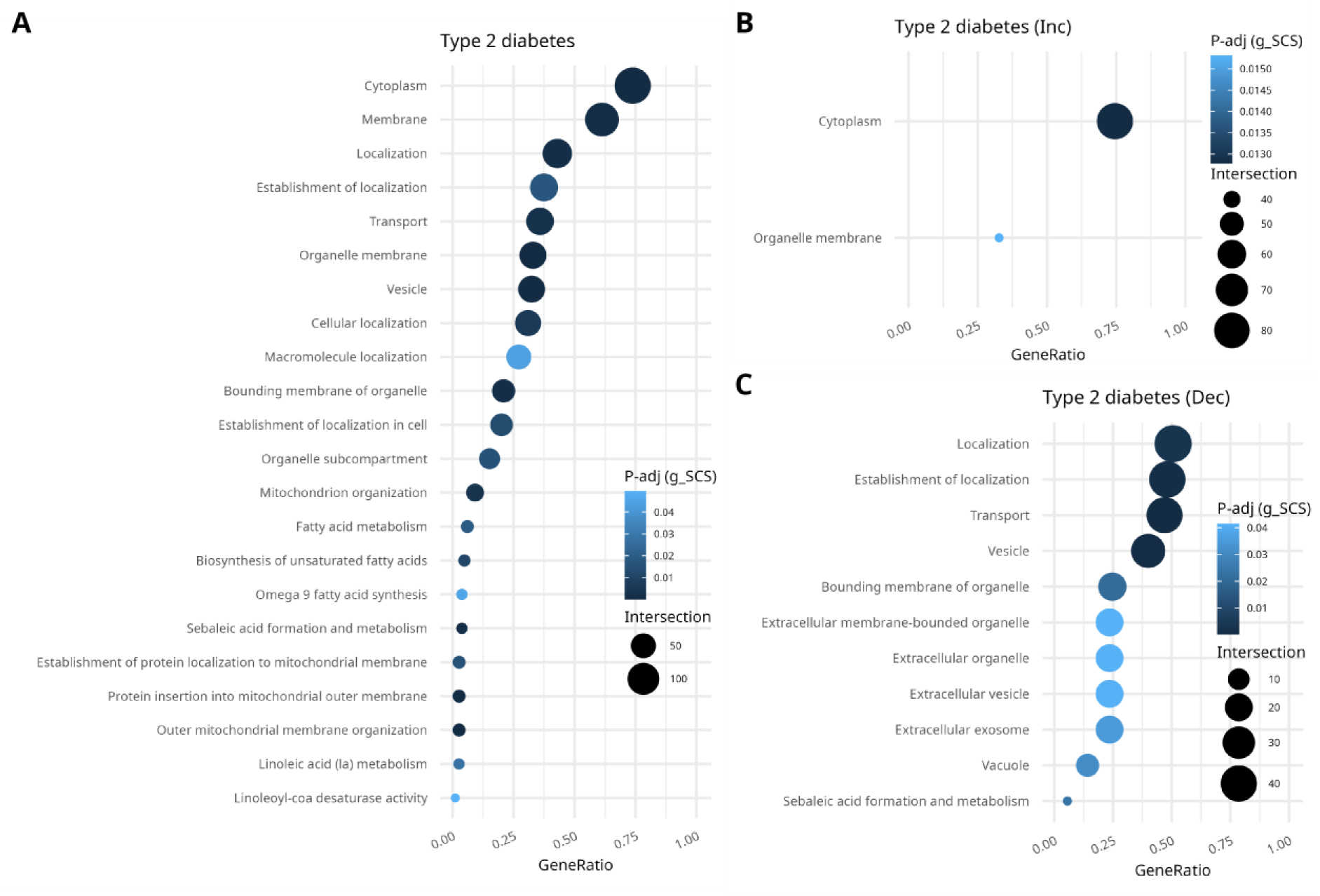
Co-localization genes are enriched in important pathways. (A) Gprofiler2 enrichment in co-localizing genes for type 2 diabetes and glycemic trait GWAS. (B) Enrichment for genes upregulated by lead risk allele. (C) Enrichment for genes downregulated by lead risk allele.

Among the type 2 diabetes genes in the vesicle pathway (Figure 5A), we identified a novel co-localization for a low minor allele frequency (5%) variant for *MYO5C* (Figure 6). The low frequency allele confers protection against type 2 diabetes and increases *MYO5C* gene expression in human islets. *MYO5C* encodes myosin type V, a molecular motor that moves along actin filaments and is involved in secretory granule trafficking (32). The lead variant rs149336329 is located in promoter and transcript regions, as well as in a human islet-specific active promoter (marked by mediator, H3K27ac and H3K4me3) (Supplementary Tables S9 and S10) near the transcription start site (Figure 6A), making it a plausible causal variant. A different lead SNP (rs3825801) is reported in (10) after fine-mapping but *coloc* co-localization was unsuccessful.

**Figure 6.**
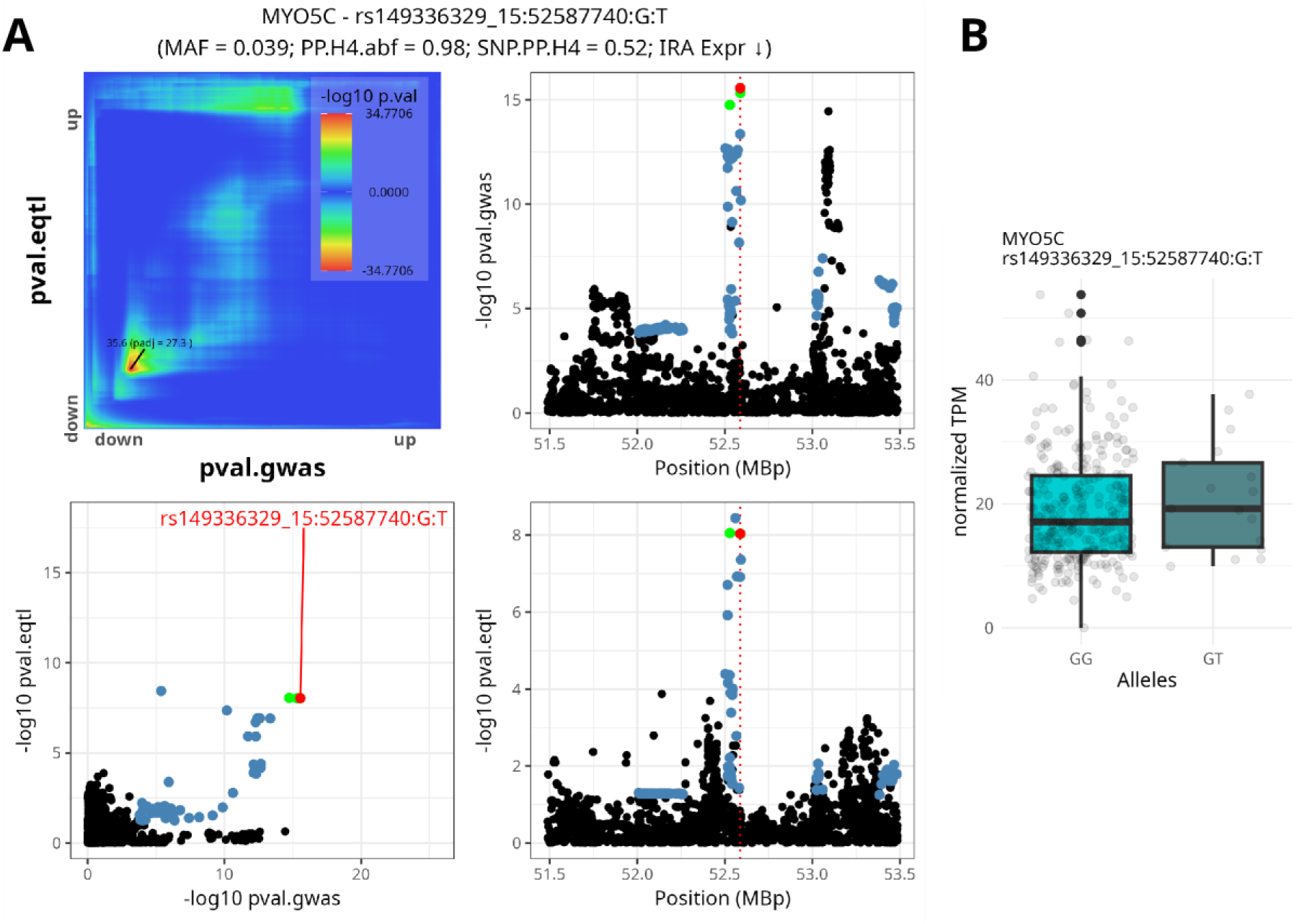
Identification of *MYO5C* as a novel type 2 diabetes gene. (A) The co-localization plots for *MYO5C*, showing the lead SNP in red, the 99% credible set in green and *RedRibbon* overlap in blue. The red dotted line is the gene transcription start site. (B) Expression plot for the co-localizing SNP. SNPs are referenced by rsids, PP.H4 abf and SNP PP.H4 are the posterior probability for *coloc* tool hypothesis 4, MAF is minor allele frequency, Increasing Risk Allele expression regulation direction; an arrow pointing down means decreased gene expression for the risk allele.

*RPL39L* stands out among the genes in our analysis, featuring a unique variant in its 99% credible set that was not detected by *coloc* alone, but identified using *colocRedRibbon* (Figure 7). The lead variant rs3887925 is noteworthy due to its presence in multiple regulatory regions, i.e. promoter flanking region, transcript, and islet specific active enhancer. RPL39L has been previously associated to type 2 diabetes in differential gene expression studies of islets from type 2 diabetic vs non-diabetic donors (27), a finding corroborated by our type 2 diabetes fGSEA enrichment analysis (Supplementary Table S1, column *fGSEA.t2d*). The risk allele T is associated with increased *RPL39L* expression (Figure 7B). *RPL39L* encodes ribosomal protein L39-like; its function in islets remains to be experimentally assessed. Interestingly, the same variant also affects *ST6GAL1* expression although *ST6GAL1* is not part of the leading edge in our type 2 diabetes *fGSEA* enrichment for human islets (Supplementary Table S11, column *fGSEA.t2d*). *RPL39L* and *ST6GAL1* are co-expressed in multiple tissues, including thyroid, prostate, ovary, artery and fibroblasts (33). Notably, *ST6GAL1* is implicated in several cellular pathways identified by our enrichment analysis, i.e. organelle sub-compartment, bounding membrane of organelle, organelle membrane, membrane, and cytoplasm (Figure 5, Supplementary Table S11).

**Figure 7.**
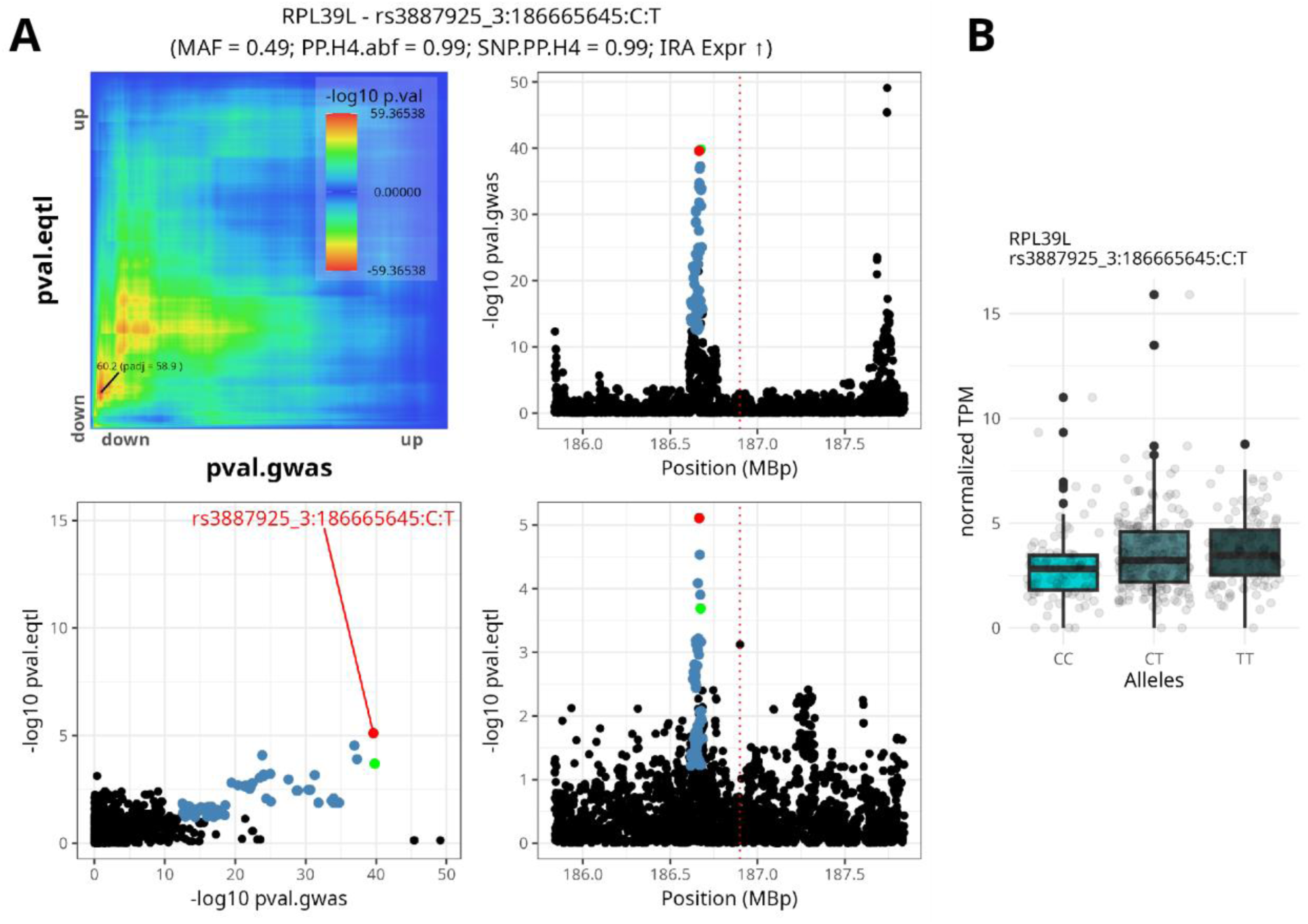
Co-localization for *RPL39L* eQTL and type 2 diabetes GWAS. (A) The co-localization plots for *RPL39L*, showing the lead SNP in red, the 99% credible set in green and *RedRibbon* overlap in blue. The red dotted line is the gene transcription start site. (B) Expression plot for the co-localizing SNP. SNPs are referenced by rsids, PP.H4 abf and SNP PP.H4 are the posterior probability for *coloc* tool hypothesis 4, MAF is minor allele frequency, IRA indicates the Increasing Risk Allele expression regulation direction; an arrow pointing up means increased gene expression for the risk allele.

*colocRedRibbon* detected 8 co-localizations for type 1 diabetes in the HLA region of chromosome 6 (Figure 2). Pathway enrichment analysis for type 1 diabetes genes did not reach significance due to the relatively limited number (n=24) of co-localizations detected.

Among the type 1 diabetes co-localizations, variant rs11666792 emerged as the lead variant for both *FUT2* and *RASIP1* (Figure 8). These genes are co-expressed in esophageal mucosa and skin (33). *RASIP1* encodes Ras interacting protein 1, that has GTPase binding and protein homodimerization activities. *FUT2*, one of the three genes that also co-localizes for type 2 diabetes (albeit with a distinct lead variant), encodes the enzyme alpha-1,2-fucosyltransferase. This enzyme is involved in the synthesis of the H antigen, the precursor of secreted ABO blood group antigens. *FUT2* has been associated with type 1 diabetes (34): a common nonsense mutation in *FUT2* abolishes secretion of ABO antigens in saliva and the gut, alters the gut microbiome and confers susceptibility to type 1 diabetes, while protecting from some viral infections (34,35). In human islets, the protective G allele of rs11666792 increases *FUT2* expression (Figure 8A). The role of FUT2 in islets is unknown, but we observed that FUT2 silencing in beta cells increased Coxsackievirus B1-induced apoptosis, suggesting that FUT2 might protect beta cells against potentially diabetogenic viruses (Figure 8C).

**Figure 8.**
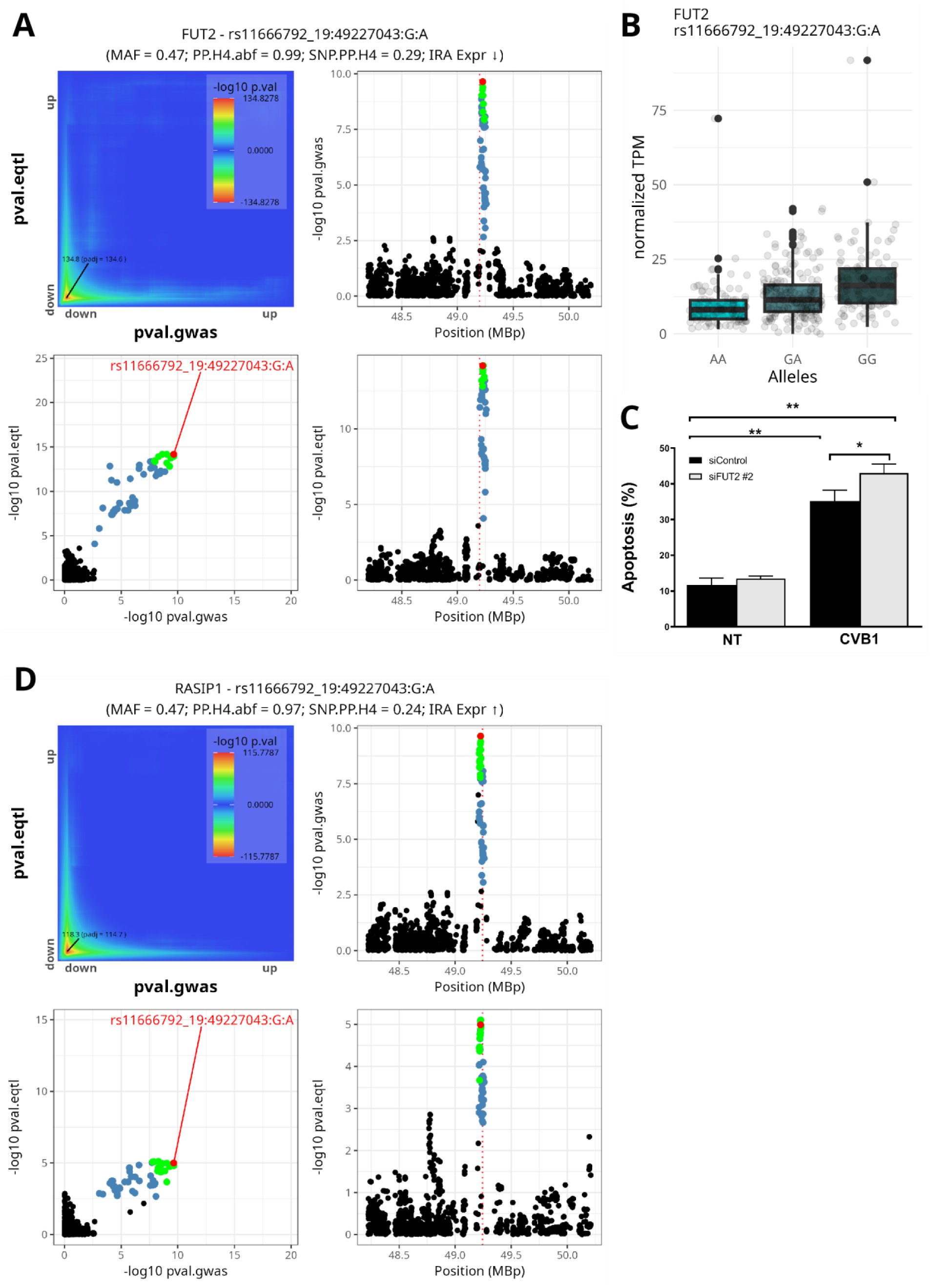
Co-localization for co-expressed *FUT2/RASIP1* eQTL and type 1 diabetes GWAS. (A) The co-localization plots for *FUT2*, showing the lead SNP in red, the 99% credible set in green and *RedRibbon* overlap in blue. The red dotted line is the gene transcription start site. (B) *FUT2* expression plot for the co-localizing SNP. (C) *FUT2* silencing in beta cells increases Coxsackievirus B1-induced apoptosis. (D) The co-localization plots for *RASIP1*. SNPs are referenced by rsids, PP.H4 abf and SNP PP.H4 are the posterior probability for *coloc* tool hypothesis 4, MAF is the minor allele frequency, IRA indicates the Increasing Risk Allele expression regulation direction; an arrow pointing up or down means increased or decreased gene expression for the risk allele, respectively.

Cathepsin H (encoded by *CTSH)*, a type 1 diabetes candidate gene (36,37), exhibits a co-localization for lead variant rs34593439, having a 0.10 minor allele frequency and a small 99% credible set (5 variants, Figure 9). The protective allele A (type 1 diabetes odds ratio = 0.84) lowers islet *CTSH* expression (Figure 9B). *CTSH* displays complex regulation in response to inflammatory stimuli. It is downregulated in the human beta cell line EndoC-βH1 upon exposure to pro-inflammatory cytokines IL-1β + IFNγ (22). The risk allele T of variant rs3825932 has been shown to reduce islet *CTSH* expression (36); this variant is in linkage desequilibrium with rs34593439. Lower methylation variability may make individuals with the protective allele potentially less sensitive to cytokines (37). The gene is also associated with celiac disease (38), type 1 narcolepsy (39), where the *A* allele of rs34593439 increases risk, and rheumatoid arthritis (40). This association with multiple autoimmune diseases makes the gene and the variant good candidates for further investigation.

**Figure 9.**
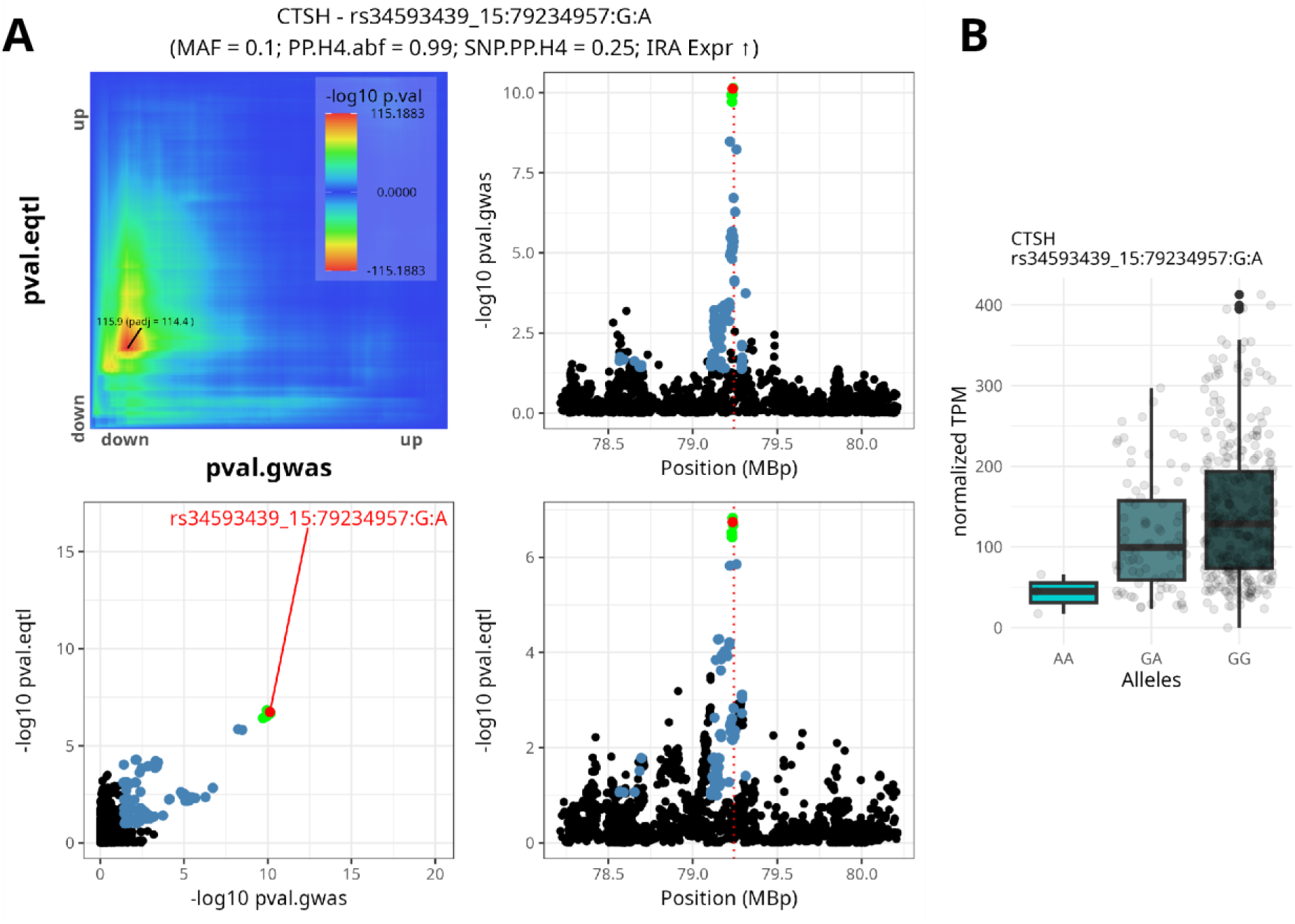
Co-localization for *CTSH* eQTL and type 1 diabetes GWAS. (A) The co-localization plots for *CTSH,* showing the lead SNP in red, the 99% credible set in green and *RedRibbon* overlap in blue. The red dotted line is the gene transcription start site. (B) Expression plot for the co-localizing SNP. SNPs are referenced by rsids, PP.H4 abf and SNP PP.H4 are the posterior probability for *coloc* tool hypothesis 4, MAF is the minor allele frequency, IRA indicates the Increasing Risk Allele expression regulation direction; an arrow pointing up means increased gene expression for the risk allele.

### Glycemic trait genes are shared with type 2 but not type 1 diabetes

Among the 48 co-localizations identified for glycemic traits, only *SULT1A2* was present among the type 1 diabetes genes. In contrast, 19 (50%) glycemic trait genes were detected for type 2 diabetes, with 11 sharing an identical lead variant. A striking example of this overlap is the single lead variant rs11708067 affecting *ADCY5* expression (41), which co-localizes for type 2 diabetes and all glycemic traits (Figure 10). The type 2 diabetes risk allele A decreases human islet *ADCY5* expression (Figure 10A-B). Consistent with its effect on type 2 diabetes risk, the A allele of rs11708067 is associated with increased HbA1c and fasting and 2-hour glucose and decreased human islet *ADCY5* expression (Figure 10C-E). Conversely, the A allele is associated with increased fasting insulin and increased *ADCY5* expression (Figure 10F). *ADCY5* encodes adenylate cyclase 5, an enzyme that converts ATP to cAMP, generating an essential amplifier of insulin secretion. Decreased *ADCY5* expression impairs glucose-induced cAMP production and insulin secretion (42). These congruent co-localization data strongly support rs11708067 as the causal variant that increases diabetes risk by decreasing *ADCY5* expression. This finding underscores the importance of ADCY5 in glucose homeostasis, highlighting how a single genetic variant can have pleiotropic effects on related traits.

**Figure 10.**
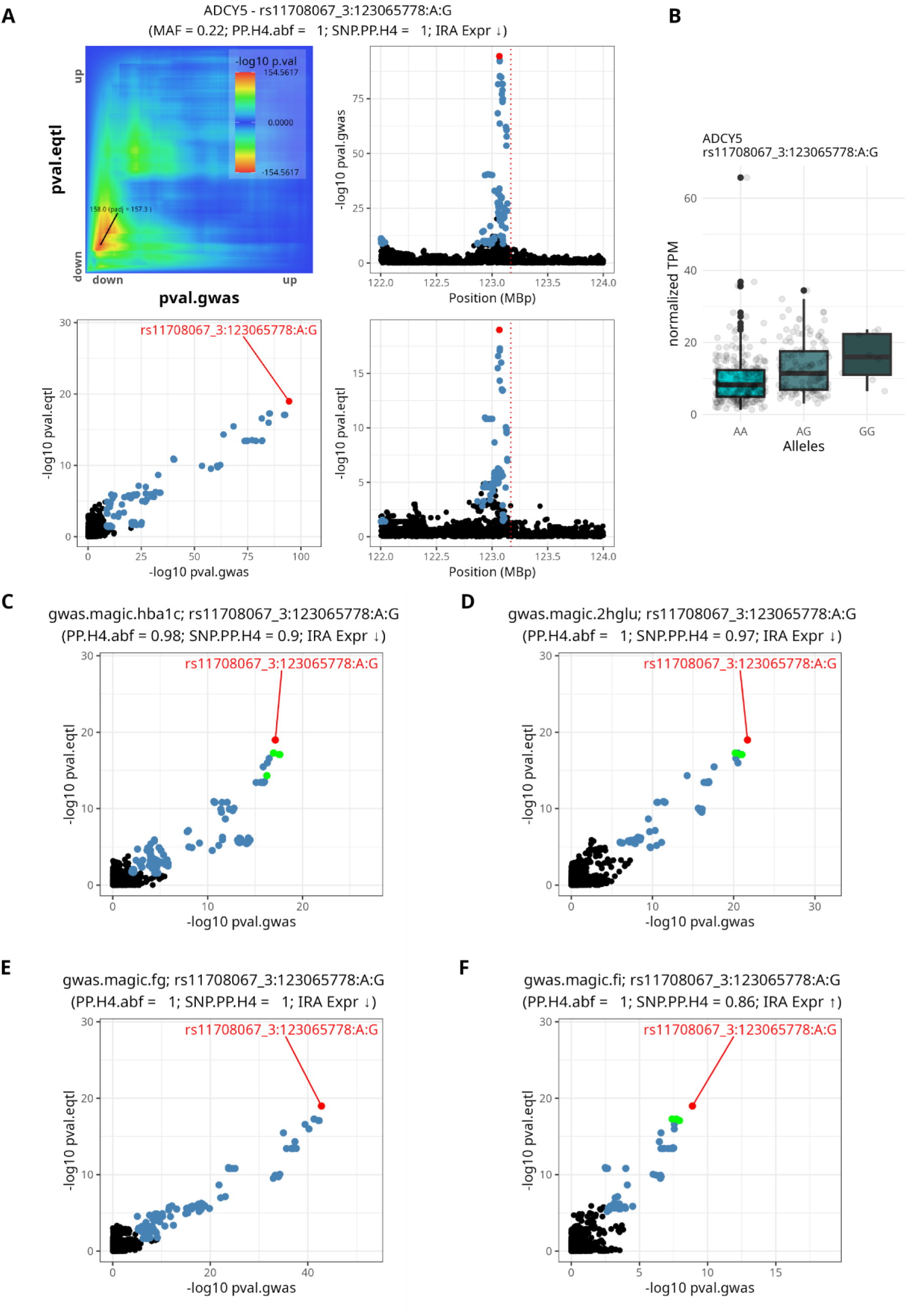
Co-localization of *ADCY5* eQTL and type 2 diabetes and glycemic trait GWAS SNPs. The co-localization plots for *ADCY5* show the same lead SNP in red, the 99% credible set in green and *RedRibbon* overlap in blue. The red dotted line is the gene transcription start site. (A) Type 2 diabetes co-localization. (B) Expression level for the alleles regulating *ADCY5*. (C-E) Co-localization for (C) glycated hemoglobin (HbA1c), (D) 2-h glucose (2hGlu), (E) fasting glucose (FG) and (F) fasting insulin (FI). SNPs are referenced by rsids, PP.H4 abf and SNP PP.H4 are the posterior probability for *coloc* tool hypothesis 4, MAF is the minor allele frequency, IRA indicates the Increasing Risk Allele expression regulation direction; an arrow pointing up or down means increased or decreased gene expression for the risk allele, respectively.

## DISCUSSION

Over the past decades, a plethora of GWAS has linked common genetic variation to complex diseases and traits. Because around 90% of all GWAS hits lie in noncoding regions, the identification of genes that mediate susceptibility is not straightforward. eQTL studies in disease-relevant tissues can connect SNPs to functional genes, but the yield of GWAS and eQTL variant co-localization studies tends to be low. This is illustrated with the >700 variants identified in diabetes GWAS, of which only around 50 have so far been linked to changes in gene expression in human pancreatic islets. Here, we significantly expanded the number of co-localizations for type 2 diabetes and identified many novel type 1 diabetes co-localizations using the novel *colocRedRibbon* pipeline. This pipeline detected 434 co-localizations corresponding to 289 genes and 344 distinct lead variants. This increase is attributed to a combination of the new computational methods, and larger, multi-ethnic and multi-trait GWAS. Without the methodological advancements of *colocRedRibbon*, we would have identified only 214 colocalizations (128 distinct gene regions), i.e. less than half of what is presently observed, highlighting the importance of these improvements in uncovering novel associations. *colocRedRibbon* filters variants or delimits regions (IQR mode, Figure 1B) to be further analyzed by a colocalization tool. We used *coloc* for subsequent steps, but any other tool may be used (e.g., *SuSiE* + *coloc* (43,44) or *smr* (*45*)).

Our analysis revealed that 88% of the lead variants are localized in regulatory regions, including those specific to pancreatic islets. For type 2 diabetes and glycemic traits, the regulated genes are involved in key pathways related to cellular localization, vesicle dynamics, mitochondrial function, fatty acid metabolism and synthesis (Figure 5). The enrichment in regulatory regions and pathways crucial for beta cell function/survival demonstrates the potential of *colocRedRibbon* to identify disease-relevant mechanisms. When comparing the detected genes with those reported as differentially expressed in islets in type 2 diabetes (27), 51 genes overlapped. This is substantial overlap, considering the relatively modest transcriptome sample size (28 type 2 diabetic and 58 non-diabetic donors) compared to the large GWAS and eQTL datasets. The overlapping genes were primarily involved in cellular localization and vesicle pathways.

Co-localizing lead variants were also found in genes involved in monogenic diabetes, such as *FOXA2* and *PCBD1.* The transcription factor FOXA2 contributes to pancreas development and islet gene expression. *FOXA2* mutations have been implicated in diabetes and congenital hyperinsulinism (46). PCBD1 acts as a cofactor of transcription factor HNF1A, which is essential for pancreatic development and function (47). These findings underscore parallels between monogenic and polygenic forms of diabetes.

The HLA region on chromosome 6 resulted in 8 co-localizations for type 1 diabetes (Figure 2). This region poses a significant challenge for co-localization analyses due to the complex structure of the linkage disequilibrium. It contains numerous immunity related genes that are under strong selective pressure (48), leading to highly intricate linkage disequilibrium patterns. For several co-localizations (e.g., *BTN2A3P*, *HCG20*), *colocRedRibbon* detected significant subsets of variants through the overlap prefiltering step. The visual representation of these co-localizations (see Data Availability) appears inconclusive, however, as the *colocRedRibbon*-identified variants are contained within stronger GWAS signals. This could be a side effect of the simplification used to assess the linkage disequilibrium in adjusted *P*-value computation, overestimating the overlap significance or, alternatively, a reflection of the method’s ability to discern multiple overlapping GWAS signals, resulting in valid co-localization. Other co-localizations in the HLA region (e.g., *BAG6*, *PSMB9*, *HLA-DQA1-AS1*) appear more convincing, with well isolated GWAS signals and the lead variant positioned close to the top of the signal. Future validation of HLA region co-localizations is required, probably using specialized tools tailored to handle the unique challenges posed by this genomic locus. Adapting and fine-tuning *colocRedRibbon* may help to address the complexities of the HLA region, but this will require a large, dedicated effort.

*colocRedRibbon* can now be applied to eQTL studies in other disease-relevant tissues, such as the immune system for type 1 diabetes (49), as well as muscle, liver and adipose tissue for type 2 diabetes and glycemic traits (50–55). It is also applicable to quantitative trait loci for protein, open chromatin and DNA methylation. The use of the same powerful pipeline across multiple tissues and quantitative trait loci will comprehensively characterize the tissue-specific genetic architecture of these polygenic diseases.

In summary, this study leveraged the novel *colocRedRibbon* pipeline to analyze a large dataset from human pancreatic islets, a tissue central in the pathogenesis of diabetes, allowing to investigate the expression regulatory variation underlying this complex disease. The innovative *colocRedRibbon* pipeline significantly enhanced detection of co-localizations of eQTL and GWAS variants compared with previous methods, refining the mapping of lead variants and uncovering numerous novel co-localizations. These results are a critical step toward understanding how diabetes-associated variants impact pancreatic islet cell function, elucidating the mechanisms linking genetic variation with diabetes and, potentially, guiding translation of GWAS hits to (personalized) diabetes therapy.

*colocRedRibbon* is a powerful new tool for integrating genetic and gene expression data to elucidate the complex interplay between genetic variation and disease risk. It is poised to expand knowledge of the genetic architecture of many complex diseases and highlight avenues for therapeutic development.

## DATA AVAILABILITY

The R package code is open to the community with a permissive license (GPL3) and available for download from GitHub https://github.com/antpiron/RedRibbon and https://github.com/antpiron/colocRedRibbon.

## SUPPLEMENTARY DATA

The eQTL dataset is from (17). The GWAS are from (11,15) and GCST90013791. The list of co-localizations is available on Zenodo via accession number #######.

## ACKNOWLEDGEMENT

We thank Piero Marchetti, University of Pisa, and his team for their essential role in providing human islet samples and data to TIGER.

## FUNDING

This work has been supported by the European Union’s Horizon 2020 research and innovation program T2DSystems under grant agreement no. 667191, the Fonds National de la Recherche Scientifique (FNRS), the Walloon Region SPW-EER Win2Wal project BetaSource, Belgium, the FWO and FRS-FNRS under the Excellence of Science (EOS) programme (Pandarome project 40007487), the Walloon Region strategic axis FRFS-WELBIO, and the Innovative Medicines Initiative 2 Joint Undertaking under grant agreement 115797 (INNODIA) and 945268 (INNODIA HARVEST). This latter Joint Undertaking received support from the Union’s Horizon 2020 research and innovation programme and the European Federation of Pharmaceutical Industries and Associations, JDRF, and The Leona M. and Harry B. Helmsley Charitable Trust. A.P. is supported by Fonds David et Alice Van Buuren, Fondation Jaumotte-Demoulin, Fondation Héger-Masson, Fondation Wiener-Anspach and FNRS. F.S. is supported by a Research Fellow FNRS fellowship. D.L.E. is supported by grants from the JDRF International (now T1D Breakthrough) (3-SRA-2022-1201-S-B and 3-SRA-2022-1201-S-B); the National Institutes of Health Human Islet Research Network Consortium on Beta Cell Death & Survival from Pancreatic β-Cell Gene Networks to Therapy (HIRN-CBDS) (grant U01 DK127786); and the National Institutes of Health NIDDK grants RO1DK126444 and RO1DK133881-01. J.M.M. is supported by American Diabetes Association grant #11-22-ICTSPM-16, NHGRI U01HG011723, the National Institute Of Diabetes And Digestive And Kidney Diseases of the National Institutes of Health under Award Number R01DK137993 and U01 DK140757, and a Medical University of Bialystok (MUB) grant from the Ministry of Science and Higher Education (Poland).

## CONFLICT OF INTEREST

None.

## Notes

### Competing Interest Statement

The authors have declared no competing interest.

### Author Declarations

The study used (or will use) ONLY openly available human data that were originally located at: - https://tiger.bsc.es/ - https://diagram-consortium.org/downloads.html - https://genetics.opentargets.org/study/GCST90013791 - https://magicinvestigators.org/downloads/

